# Perceived benefits and challenges of school feeding programs in Addis Ababa, Ethiopia: A qualitative study

**DOI:** 10.1101/2023.10.08.23296723

**Authors:** Yihalem Tamiru Semegn, Samson Gebremedhin Gebreselassie, Afework Mulugeta Bezabih, Abebe Ayelign Beyene, Elyas Melaku Mazengia

**Author notes:** Corresponding author E-mail:* *(YT).

## Abstract

**Background:** School feeding programs aim to reduce children’s immediate hunger, and improve health and education outcomes. However, there is limited data on the perceived benefits and challenges of the school feeding program in Addis Ababa. This study aimed to explore teachers’, students’, and parents’ perceptions of the school feeding program in Addis Ababa, Ethiopia.

**Methods:** A qualitative study was conducted in Addis Ababa from April 10 to May 26, 2023. In-depth interviews with children, school directors, and focus groups with parents in 20 randomly chosen public primary schools were used to collect data. A total of 88 participants were included in the study: 20 students (6th and 7th grade), 48 parents, and 20 school directors. The non-probability sampling technique was used to select participants. All data collected in the local language and interviews were audio recorded, transcribed verbatim, and translated into English. The data were coded using ATLAS-TI version 9.1.3.0 software, and thematic analysis was done.

**Results:** The findings were presented as two major themes and eight sub-themes. 1) "Perceived benefits of school feeding programs," which comprised of four sub-themes, “improve educational and health outcomes, "reduce the socioeconomic burden of the family," "student behavior change" and "decreased social stigma and increased social integrity" sub-theme. 2) "Perceived barriers to SFPs gaps " include "poor market linkage between fostering mothers and producers," "poor infrastructure," underpayment of workers" and "increased sense of dependency."

**Conclusions:** The key perceived advantages of school feeding programs are improved students’ educational, health, behavior, and social integrity. Reduce the socioeconomic burden of the family. In particular, identified barriers and challenges include a lack of market linkage, poor infrastructure, and an emergent sense of dependency. Developing market connections, improving school facilities, raising public awareness of the risks associated with reliance on the school food program, and emphasizing the need for additional research to quantify potential benefits and obstacles of school feeding programs.

## Introduction

The school feeding program is one of the world’s largest and most widespread social safety nets, benefiting 418 million children worldwide (1). An estimated 66 million primary school children attend school hungry, struggle to learn, have poor concentration, and have little interest in learning (2–6), and 23 million hungry children in Africa (6). More than 121 million school-aged children are still not enrolled, with two-thirds of them being girls living in rural areas in the world’s most vulnerable regions (7).

The World Bank and the World Food Programme published a joint review of SFPs in 2009 (8) reinforcing the rationale and objectives of SFPs. The three main goals identified were to provide safety nets for families to absorb social and economic shocks, to improve school-aged children’s education and scholastic performance, and to improve children’s nutrition and health status (8).

School feeding programs (SFPs) are widely regarded as a game-changing option for improving food availability and education, as well as a prominent and innovative vehicle for addressing multiple Sustainable Development Goals (SDGs) outcomes (1,9). School feeding programs help school-age children and adolescents develop physically, mentally, and emotionally, especially in low-and middle-income nations (6).

In 1994, the Ethiopian government and the World Food Programme (WFP) launched the school feeding program to address problems identified on the ground through School Feeding Programmes (SFPs) (10). Malnutrition impairs the academic performance of schoolchildren from low-income families (11). A 2015 study in Ethiopia found that malnutrition affected 31% of schoolchildren (19.6% stunted, 15.9% underweight, and 14.0% wasted) (12). Furthermore, studies have shown that adolescent girls and boys aged 15 to 19 years are prone to chronic energy deficiency (11,13). Attending classes while hungry hurts children’s and adolescents’ ability to learn, thrive, and reach their full potential (6,14).

The Addis Ababa school feeding program was designed to alleviate hunger among poor schoolchildren, improve attendance, reduce dropout and repeat rates, and, ultimately, improve student academic performance (11,15). In February 2019, the Addis Ababa City Administration launched a broader school feeding program to serve all pre-primary and primary school students, though charitable societies and other stakeholders had previously provided school meals in some schools (11,15).

Recognizing this fundamental fact, Ethiopia’s government has made every effort to ensure that all of the country’s children have access to education. Several donors and national governments from both developed and developing countries have also invested millions of dollars in school feeding programs (10,16). Despite the attention and resources devoted to school feeding programs, little rigorous evidence exists to support these investments (16), and no adequate research on SFP has been conducted in our country to date (10,17,18), and there is a lack of studies that directly address the unknown perceptions of parents, teachers, and students towards the school food program (19). As a result, the purpose of this study was to investigate the perception of parents, teachers, and students towards the school feeding program in public primary schools from Addis Ababa, Ethiopia.

## Materials and Method

### Study setting and period

This research was carried out in Addis Ababa, the capital city of Ethiopia (20), from April 10 to May 26, 2023. It is the country’s largest city and plays an important political, economic, and symbolic role in Ethiopia (20). The city was divided into 11 sub-cities and 116 districts, and according to the 2007 population census, there were a total of 3,384,569 populations and the population will exceed 5 million in 2036 (20). There are 349,696 primary school pupils in grades 1 to 8 (21), and the SFPs have now been implemented in all 264 public primary schools located in all sub-cities, with 638,857 students benefiting from the SFP (21).

### Study design

This study employed a qualitative approach because it relies on the opinion of individuals; and asks broad, general questions and data collection consists of a large of words or texts. To examine the perceived benefits and challenges of the school feeding program in the primary schools of Addis Ababa city. For this study, in-depth interviews were used as other methods, and a case study approach was used. The SFP in Addis Ababa was taken as a case because it is the pioneer program, which is sponsored by the government and delivered to all public primary and pre-primary schools in Addis Ababa. A case study is a qualitative research method that enables an in-depth examination of a phenomenon or a program using a variety of data sources in its natural context (22,23) The constructivist paradigm was used to analyze the data.

### Study participants

The target population was school directors, parents of students, and students in grades 6 and 7 who were recruited purposively in the current study. Those who met all inclusion criteria but declined to participate were not included in the study. The study only includes school feeding program beneficiary students and their parents.

### Sampling process

The participants in the study were chosen using a multi-stage sampling procedure. Three rounds of sampling were used to choose the final research subjects. In the first step, five of the eleven sub-cities (or 50% of the sub-cities) were randomly chosen using a lottery technique. The second phase was providing a list of each sub-city’s public primary schools. Next, four elementary schools from each sub-city were chosen at random.

The study population consisted of adolescents between 12 to 19 years of age. Study participants attended grades 6 and 7 in government primary schools of Addis Ababa. While selecting schools in each sub-city, a random sampling technique was employed. Depending on the study’s design, the population’s diversity, and the depth of the data, many academics advise using a range of research participant sizes. For example, Lincoln and Guba advise 12–20 participants for studies based on interviews (24). Taking into account these factors, a total of 20 participants were purposively selected (1 with good academic performance, 1 with average academic performance, and 1 with poor academic performance) in each school including both (grade 6 and grade 7). After briefing school principals about the objectives of the study, they assisted in the selection of students based on their academic performance from grades 6 and 7. This method of sampling is employed to obtain a more diverse and representative sample (25).

After interviewing 14 participants, the interviews were stopped due to data saturation, which was the point at which further interviews produced no new data (26). Key informant interviews (KIIs) were also conducted with school principals or their deputies on their perception of the perceived benefits and challenges of school feeding programs. In total, this study was conducted on 20 adolescents, 20 primary school directors, and 5 FGDs (48 parents of students) who participated in the study. The lead investigator, who has experience gathering qualitative data and has taken advanced qualitative research methodologies, interviewed 20 school directors in-depth.

### Data collection tool and data collection process

Data were collected through an in-depth interview (IDI) using semi-structured interviews, and focused group discussions (FGDs) guide. Before being used for the actual data collection, these guides were produced by reviewing various literature and having it evaluated by three experts in the field. A semi-structured interview guide was used and the guide was prepared in the English Language following the review of various literature (23,27,28). All interviews were conducted in Amharic (the national language) and were audio-recorded with the consent of the study participants. The IDI and FGDs have focused on how people view the school feeding program and how they perceive its implementation. The data collection process through IDI was carried out by experienced research assistants, three females and two males and all five have master’s in public health and nutrition, who conducted the interviews after attending a two-day training workshop.

During the data collection period, the corresponding author supervises the whole process at the site and provides regular feedback and corrective actions. The interview was conducted in a quiet and stable condition to avoid the diversion of attention until information saturation was obtained. Interviews with all respondents were conducted face to face. In-depth interviews were conducted in the office of the school directors and lasted between 40 and 60 minutes.

Five focus group discussions with parents of students took place in a private conference room and lasted between 60 and 90 minutes. As a starting point for our investigation into the enablers and impediments to SFP in Addis Ababa city, we also used the supervision report developed by the Addis Ababa City Administration School Feeding Agency and education bureau supervision monitoring and support findings report on the performance of SFP activities in the 2022–2023 academic year.

The data were collected using pre-tested semi-structured in-depth interview guides. The recruited five data collectors who owned a master’s degree in public health and nutrition had practical experiences in the whole process of a qualitative inquiry. They were trained for two days on the objectives of the study, data collection techniques, and local and international ethical principles. The lead researcher moderated and conducted focused group discussions (FGDs) with one assistant in the conference room. Interviews with school directors took place at their offices in the morning. We picked the morning hours to minimize the impact of teachers’ work-related fatigue on the quality of the data. To reduce socially desirable responses to interview questions, all of the student interviews took place in a separate room with just the interviewee present. After reaching a saturation level, interviews and FGDs were stopped.

### Focused group discussion participants

The study’s objectives and methodology were explained to school principals both in person and via informational correspondence. They were required to explain the goals of the study to all parents and request their participation. In five participating public primary schools between April and May 2023, five focus groups with 8–10 people each were convened (23). Parents of students participated in each of the five focus group discussions (n = 48; 10, 10, 9, 9, and 10 participants).

### Focus group methodology

A semi-structured FGD guide was applied to gather information from parents about the perceived benefits and challenges of school feeding programs. The FGD used a guide including questions to explore the perception and opinion towards the school feeding program. In addition, questions regarding potential impediments and enablers to the implementation of school food programs were posed to the FGD participants. Two researchers were present during the focus groups: the moderator, who asked the questions and oversaw the discussion, and the assistant moderator, who was in charge of the recording equipment and recorded the responses from the participants.

The assistant moderator’s notes were used when necessary during the transcription of all group conversations, which were all recorded. The moderator described the study’s purpose and objectives before starting the discussion. Each focus group lasted 60 to 90 minutes and parents provided their consent. The study procedures were officially approved by the IRB.

### Focus group questions

Discussion guides were developed based on the topics. The discussion guides were developed during meetings of the multidisciplinary research team, consisting of nutritionists, behavioral change communication, and public health experts. Their formulation of the discussion guide was guided by the appropriate literature, research objectives, and other topics related to the program’s implementation. Moreover, the first focus group with parents served as a pilot group. Following pilots, no structural changes were necessary, and data were collected. For this reason, the pilot focus groups were also included as actual data of the study.

Participants answered ten questions, covering the following topics: (i) how do parents view the impact of the school feeding program on their children’s education and health outcomes? (ii) How do parents view the impact of the school feeding program on socioeconomic issues? (iii) What is the best way to improve the school feeding program? (iv) any changes seen in their children following the school meal program. The moderator was careful not to steer the questions or express any form of acceptance or criticism of the participants’ answers, and participants were encouraged to share their own opinions.

### Key informant interviews

Following the FGDs, 20 key informant face-to-face interviews were conducted to gain deeper insight into the opinions and experiences of a school director on the implementation of the school feeding program. Participants responded to 10 questions on the following topics: (i) how do school directors feel about the school feeding program’s implementation about the socioeconomic concerns of students and parents? (ii) What do school directors see as obstacles to and enablers of the implementation? (iii) How do the school directors see the school feeding program’s impact on improving the academic and health outcomes for the benefited students? (iv) What suggestions do the school directors have for the program’s further improvement?

### Student interviews

To examine students’ perceptions and opinions on the school feeding program, In-depth qualitative interviews were undertaken with students from selected public schools. Based on the degree of saturation a total of fourteen children were among the 20 study participants. Participants responded to 8 questions on the following topics: (i) What impact do students believe the school feeding program has had on their academic success and health? (ii) what problems have you noticed with the program’s implementation? (iii) How do the students see the impact of the school lunch program on family income? (iii) what suggestions do the kids have for enhancing the school lunch program?. In-depth interviews were conducted in the separate class and lasted between 30 and 45 minutes.

### Data quality assurance

Appropriate note-taking and abstraction were carried out to maintain the quality of the data. The credibility and dependability of the data were maintained by continual follow-up aside from data triangulation in time, person, and place.

### Data analysis

The collection and analysis of data started simultaneously. For analysis, the verbatim Amharic transcriptions of the audio files were translated into English after being translated into Amharic. The method of theme analysis recommended by Braun and Clark (29), was used. It started with a careful reading of the transcripts using the resilience framework’s theoretical lens (29,30). To fully understand the data and generate initial codes, two authors independently read and reread the transcripts. After that, thematic analysis was carried out by categorizing codes with related concepts into groups, and groups of categories into themes. Atlas. it was used to analyze all the data to create a final coding scheme based on emergent theme identification. Themes were subsequently adjusted and new themes were added as they emerged from the data. Through a series of discussions, these codes were refined, mapped, and organized into themes which were further redefined through several iterations until the research team reached consensus on the meaning and interpretation of the thematic areas. To validate deeper contextual understandings, the draft findings were also shared with stakeholders and academics who are culturally competent. Following an in-depth discussion among all the authors, the analysis concluded.

### Trustworthiness and rigor

Different measures were taken to ensure the trustworthiness of these findings, including participant triangulation (data were gathered from students, parents, and teachers), method triangulation (in-depth interviews, FGDs, and document reviews), and extended engagement to build rapport and trust among participants. Expert-reviewed interviews and FGD guides were employed.

### Ethical considerations

Ethical approval for this study was obtained from the Institutional Review Board of the College of Natural and Computational Sciences (CNS-IRB), Addis Ababa University, with the reference number CNCSDO/623/15/2023. Before approaching study participants, the necessary permission was sought from the Addis Ababa Education Bureau and all selected sub-cities. All interviews and focus group discussions were carried out in the privacy of the study participants; necessary precautions were taken to avoid intruders or any situation that could make the participants uncomfortable. Verbal informed consent was obtained before starting each interview which included permission to use audio recorders. Data were then kept securely and only accessible to the research team members. Before the study began, the guardians or parents of the students as well as the school’s director gave their informed consent. Students gave their informed assent and voluntarily took part in the study. All data collectors, supervisors, and investigators adhered to strict confidentiality standards by using code numbers rather than names and locking up all of the data.

## Results

### Characteristics of the study participants

Parents, students, and school directors from 20 public primary schools in Addis Ababa, Ethiopia, were the study’s target population. The total participants were 88 (48 student parents, 20 students, and 20 school directors) participated. Every parent’s child participated in the school feeding program. The parents were categorized into 5 focus group discussions.

The educational status of the study participants only two of the school teachers had advanced degrees, while 18 of them had a BA or BSc degree. Forty of the parents of students in the groups had formal education up to the certificate level, with 23 having completed the 9th through 12th grades, 12 having completed the 4th to 6th grades, and the remaining 5 having certificates. The other eight participants had no formal education but could read and write.

About the employment status of the parents of students, 30 work for private or public organizations at various levels, while the remaining 18 are housewives. Regarding their ages, there were 27 between the ages of 25 and 36 and twenty-one parents between the ages of 38 and 45. Twenty 6th and 7th-grade students were selected for in-depth qualitative interviews. In this group, there were 13 female students and 7 male students. 11 of the students were between the ages of 12 and 14, 7 were between the ages of 15 and 17, and the final two were between the ages of 18 and 19. (Table 1).

**Table 1:** Socio-demographic characteristics of the study participants from Addis Ababa, Ethiopia, 2023 (n = 88).

| Categories | Age |  | Sex |  | Educational level |  | Occupation | Total |
| --- | --- | --- | --- | --- | --- | --- | --- | --- |
|  | Min | Max | Male | Female | Min | Max |  |  |
| Students | 12 | 19 | 7 | 13 | 6<sup>th</sup> | 7<sup>th</sup> | Students | 20 |
| Teachers/ SFP focal | 30 | 38 | 1 | 1 | BA | BA | Teachers | 2 |
| School directors | 25 | 52 | 7 | 11 | BA | MA | Teachers | 18 |
| Parents | 25 | 45 | 0 | 48 | Read and write | Diploma | Housewives, civil servants, private employees | 48 |
| Total |  |  |  |  |  |  |  | 88 |

### Theme 1: Perceived benefits of the school feeding program

The following themes were identified for the perceived benefits of school feeding programs in Addis Ababa, Ethiopia.

#### Increases students’ school enrolment and academic performance

The majority of the participants of this study explained that the school feeding program has improved students’ school enrolment and academic performance through the reduction of hunger and health problems. They also believed that the introduction of a school feeding program reduced the children’s labor abuse and enabled them to stay at school and focus on their studies as explained by the following quotes from the school director and SFP focal person.

The majority of study participants explained that the school feeding program has increased the academic achievement of students and school enrollment by reducing hunger and health issues. According to the following quotes from the school director, students, and parents, they additionally believed that the implementation of the school feeding program reduced the children’s exposure to child labor abuse and allowed them to attend class and focus on their studies.

> *“Since the introduction of the school feeding program, the number of students falling asleep in the classroom during school hours and falling sick due to hunger has decreased. This helps students to attend their studies properly without any dropout.”(School director)*
>
> *“Many students were hungry before the implementation of the school feeding program, and they were also at risk of contracting food poisoning and other food-borne illnesses by consuming leftovers outside the school. As a result of missing numerous days of school, it has a detrimental effect on their academic achievement as they are missing several days of school.” (School director)*
>
> *“Unnecessary labor exploitation was also happening to students. Instead of coming to school, students were engaged in daily labor to earn a living.”(School director)*
>
> *“Students do not have to worry about the food they are eating, so they are happy to learn their lessons, and helps them to focus on their academic only.” (Student)*

> *“Since the introduction of school meals, many students’ motivation in learning has improved, and students now actively resist being asked by their parents to miss school to work.” (Student)*
>
> *“Had poor academic performance and no interest in learning before the school feeding program, but today he is much more interested in learning and his academic performance has considerably improved.” (Student)*
>
> *“Many parents of students find themselves unable to purchase whatever their children require for school due to the high cost of living. Parents are now much more motivated to send their children to school since the feeding program was started.” (FGD participant)*

#### It decreases the socioeconomic burden on the family

The majority of participants concurred that the school feeding program lessened the financial strain on students’ families as well as the stress placed on them when students fall unconscious at school because of hunger. They also emphasized the possibility of spillover effects, in which one child’s utilization of a resource by the family may benefit another. Many participants brought up the issue that the fact that mothers of students cook food for students at school gave them job opportunities and allowed them to make extra money.

> “*The school feeding program has reduced the economic and living burden of the parents of students and has also created employment opportunities for the parents of students.”(School director)*
>
> *“Before the start of the school feeding program, many parents of students were concerned about what their children eat for breakfast and lunch and the food that is prepared for them, and the economic capacity of most parents makes it difficult to meet the basic needs of their children.”(School director)*

#### Change in students’ behavior

The majority of participants believed that students had improved behavior as a result of the school feeding program. Before the implementation of SFP, students reported used to climb over the school fence to look for food when they became hungry. Additionally, they explained that when students go looking for food, they get subjected to gender-based violence and substance abuse.

> *“Before the school feeding program was introduced, female students were often subjected to sexual harassment and assault when they skipped school to get food because they were hungry. However, after the introduction of the school feeding program, the problem has been greatly reduced.”(School director)*
>
> *“Before the school feeding program began, some students were exposed to various drugs, including khat and shisha. However, since the school feeding program was introduced, students are now safer than ever as they do not leave campus.”(School director)*

Some participants also agreed that SFP had decreased student misconduct, such as stealing money from their parents, skipping class during the school day, and fighting with the school’s security personnel. However, SFP has since helped generate the number of students who are disciplined.

> *“Before the school feeding program started, some students used to steal money from their parents to buy bread and various sweets and spend the stolen money. And it was causing them to grow up destructive rather than causing inappropriate behavior problems, but after the school feeding program was started, the problem was greatly reduced.” (School director)*.
>
> *“Before the implementation of the school food program, there were numerous behavioral issues among students, and occasionally, students would fight with the school security staff for various reasons. However, now these issues have significantly decreased.” (School director)*

> *“The school feeding program service used to be available only to the poor students, but now it is available to all students, without exception. It boosts the students’ morale, increases their interest in learning, and helps them study calmly in class." (FGD participant)*
>
> *“The safety of students has arguably been enhanced by the school feeding program in several ways, including the fact that they are less likely to contract illnesses and that their health has improved. As a result of the implementation of the school feeding program, the problem of many students being exposed to unnecessary risks by leaving the school grounds has significantly decreased.” (School director)*

#### Decreased social stigma and increased social integrity

Before the start of the school feeding program, only students from low-income families were eligible for the school feeding program, which subjected students to discrimination, as was commonly mentioned by some participants. Furthermore, the majority of participants also described that the SFP increased unity and social integrity and decreased discrimination among students when they eat together regardless of their families’ economic status.

> *“The school food program shields the students from psychological trauma and humiliation. Before the implementation of the school feeding program, a large number of students did not bring food to school, but now all students share the same meal.” (School director)*
>
> *“The school feeding program not only makes unhealthy competition among students healthy but also reduces perceived discrimination and helps students to focus on their studies by providing equal access to services for all students.” (FGD participant)*

### Theme 2: Perceived barriers to a school feeding program in Addis Ababa City

Regarding the challenges facing the city of Addis Ababa’s school feeding program, four themes were found. These included a lack of market connections between foster mothers and producers, poor infrastructure, insufficient pay for the food handlers, and a growing sense of dependence.

#### The poor market linkage between fostering mothers and producers

One of the main barriers to effective SFP was the lack of connections or linkage between foster mothers and suppliers of market items and agricultural products. Some participants indicated that the SFP’s inability to obtain essential supplies like tef, flour, oil, sugar, bread, and culinary items is impeding its effectiveness of the SFP. The participants suggested that this issue may be resolved by establishing a network with farmers, suppliers of market goods, and foster mothers.

> *“Concerning the school feeding program for students, supply and distribution of resources to foster mothers with consumers on the following resources: there is a link between sugar, oil, flour, and the sheger bread association, but it is insufficient because it does not account for the quantity and number of students.” (FGD participant)*

#### Poor infrastructure

Most participants agreed that schools lacked the necessary infrastructure to implement an efficient school feeding program. Facilities including the lack of a standard dining room, kitchen, chair, and table, as well as the lack of water and electricity supply, were frequently mentioned.

> *“Frequent water and electricity outages in our schools, as well as inflation and lack of supply, have had a major impact on the implementation of the school feeding program.”(School director)*
>
> *“About the school feeding program for students, the kitchens used for cooking in most schools are made of tin, so they do not have enough air circulation and their width is narrow, so it is difficult for the mothers who prepare the food to do their work properly.” (FGD participant)*

#### Underpayment of workers

The majority of participants mentioned underpaying mothers who prepare and serve food to students as the other barrier. Nearly the majority of the participants concurred that the present budget for mothers does not account for inflation. The other issue is the inadequate funding for the school lunch program.

> *“At our school, the major issue with the school feeding program is that the compensation given to those who serve food to students doesn’t account for inflation or the actual cost of living. For instance, the cost of breakfast and lunch for one student is currently 20 Ethiopian Birr. This quantity of money puts unneeded strain on food handlers and demotivates them at work because it doesn’t account for the current inflation.” (School director)*
>
> *“It is crucial to pay attention to the cost of food handlers’ salaries while discussing the school feeding program for students. One student only has to pay 20 birr every day in tuition. A market assessment study should be carried out to determine the compensation to be paid to food handlers who prepare meals for students in consideration of the pressures of the existing cost of living and the rise in the price of supplies. A payment increase is required.” (FGD participant)*

Many participants also remarked that parental leave without pay and the interruption of payment for food handlers during the summer months make it highly challenging for food handlers to support their children.

> *“The problem of not paying the mothers who prepare food for the students after June 30 when the school is closed. Along with this, breadwinner mothers are exposed to problems as they will not have monthly income until the school opens.” (FGD participant)*
>
> *“Mothers who prepare meals for students due to maternity leave are not paid like other workers when they leave school, and also the problem of not having an independent payment system.” (FGD participant)*
>
> *“Another challenge of program implementation is the insufficient allocated budget for a meal and the high cost of food items “ (School director)*

#### Increased sense of dependency

Some participants mentioned fear that SFP is making students’ families feel dependent on them and that students are prioritizing food before education. Some teachers also brought up the concern that students spend too much time eating at nearby cafes and not enough time studying.

> *“School feeding programs are beneficial for students. However, it would be preferable if it just applied to low-income parents of students. The school feeding program has unnecessarily raised dependence on parents of students because it includes participants from various socioeconomic backgrounds, including both pupils and parents. They are no longer able to take on responsibility or handle problems alone, and as a result, they develop a dependency mentality. “ (School director)*
>
> *“Some students give a lower priority to their education and attend school rather than eat breakfast and lunch from the school feeding program. Since the majority of students come from poor families with lower incomes. “ (Student)*.

## Discussion

The present study explored the perceived benefits and challenges of school feeding programs in Addis Ababa city using the case study design. Increments in school enrollment and academic performance, a decrease of the socio-economic burden of students’ families, decrement of social stigma and improvement of social integrity between students, and a change in students’ behavior were themes identified as perceived benefits of SFP. On the other hand, the absence of market linkage between fostering mothers, suppliers, and producers, poor infrastructure, underpayment of workers, and a sense of dependency were themes identified as key challenges for effective and efficient SFP implementation in Addis Ababa City, Ethiopia.

According to this study, the SFP is improving student enrollment, lowering the dropout rate, and enhancing academic achievement by reducing child labor abuse and hunger. This result is consistent with a quantitative study conducted in Addis Ababa, Ethiopia (31), which revealed a favorable relationship between academic achievement and SFP.

This result is consistent with a review (32) of qualitative studies that were carried out in Niger (33), and Nigeria which which looked at how SFP was considered to affect students’ enrollment, completion, and academic success (34). Therefore, strengthening the SFP could be a crucial intervention to enhance students’ academic performance and improve the quality of their education.

The other perceived benefit of SFP in Addis Ababa was the reduction of socio-economic burden on students’ families. Low-income families found it difficult to provide breakfast and lunch for their children, and when students were hungry and fell unconscious families encountered anxiety and worry. However, the SFP helped them alleviate the socioeconomic pressure on families, particularly those with lower income levels.

This finding is further supported by a report from a systematic review conducted to determine the facilitators and barriers of SFP (32), as well as a study carried out in the Sidama region of Ethiopia (35). This finding was also supported by a study conducted in Tennessee, USA, which revealed that school lunch programs reduced family stress related to both time and financial resources for grocery shopping, preparing, and packing food for their children (36). SFP therefore has the potential to serve as an essential strategy for improving the standard of living for families, especially those with low income.

According to the present study, one of the perceived benefits of SFP was a reduction of social stigma and an increment of social integrity between students. Before the endorsement of this SFP program at all public primary schools in Addis Ababa, there was a school feeding program only for students from the poorest family, and this approach had a social stigma, but now there is no eligibility criteria and every student is served breakfast and lunch at school regardless of the financial status of their parents. This approach reduced the stigma and increased social integrity among students and this finding is supported by evidence from a qualitative study done in India (37) and a qualitative systematic review, which reported that SFP increases social interaction and integrity of students (32).

SFP contributes to reducing poverty both directly and indirectly through improving community wellbeing (33). Additionally, this study discovered that the implementation of SFP decreased children’s inappropriate behaviors, such as leaping over the school gate when they were hungry, chewing chat, and being exposed to sexual abuse. This finding is confirmed by the study’s findings from Niger, which gave SFP’s role in reducing student misconduct such as theft, drug usage, sexual assault, and delinquency a lower rating (33). This suggests that SFP could be applied as a strategy for generating a disciplined and productive generation in the future.

Ethiopian school feeding policy made clear that the government is responsible for providing schools with basic facilities that are appropriate for the local environment (38). However, this study indicated that inadequate facilities, such as those in the kitchen, dining room, water supply, and electric supply, were among the main obstacles to the implementation of SFP. Qualitative research in the Sidama region of Ethiopia (35), Nigeria (34), and Uganda (39) as well as a qualitative systematic review undertaken globally (32), all provide credence to this finding. Therefore, building basic facilities as well as improving the water and electric supply could positively impact the success and sustainability of SFP in settings with scarce resources, such as Ethiopia.

This study also indicated that underpayment of workers under SFP, absence of payment for workers in the summer season, and budget that doesn’t consider inflation were other challenges to the effective implementation of SFP. This finding is supported by evidence from a qualitative systematic review in a global context (32) and a qualitative study conducted in Addis Ababa (40). These studies reported that the absence of a budget that considers inflation was a challenge for the smooth implementation of SFP. Therefore, this program should give due emphasis on adjusting the budget according to the living cost of the country.

The other major obstacle to effective SFP in the current study was the lack of networking between producers, suppliers, and the school feeding program. This resulted in a shortage of numerous inputs for the SFP, and this finding is supported by a global review of SFP (32). As a result, the improvement of the food supply chain should receive appropriate attention, as indicated in the school feeding policy (38).

According to the Ethiopian school feeding policy (38), effective and sustainable SFP needs cross-sectoral cooperation, and the local community is expected to oversee SFP implementation and allocation of resources with a sense of ownership. The SFP was, however, found to be developing a sense of dependency among the parents of the students, according to one study. Thus, to maximize the sense of ownership and minimize the sense of dependency on it, as already mentioned in the national school feeding policy, program implementers should pay appropriate attention to engaging parents in particular and the local community in general in the process of planning, implementing, and evaluating this program (38).

### Strengths and limitations

One of the strengths of this study is the school-based approach, which provided rich insight into perceptions towards the school feeding program In addition, interview facilitators were well trained, equipped with previous qualitative research experience, familiar with the community, and fluent in the local language. Relationships with the communities were established before data collection by approaching the primary school directors for prior permission. Analysis was conducted as a team with the use of multiple researchers. As with any research, there are several limitations to these findings the results of this study cannot be generalized because it is a qualitative study without a representative sample. This study would have benefited more from a mixed-method approach to quantify the size of each perceived advantage and challenge associated with SFP so that planners and policymakers may more easily prioritize problems.

## Conclusion

The key perceived advantages of SFP in Addis Ababa, Ethiopia, included an increase in student enrollment, attendance, and academic performance; a decrease in the socioeconomic burden on students’ families; an improvement in students’ behavior; a reduction in social stigma; and an increase in social integrity among students. The key obstacles to the successful implementation of SFP in Addis Ababa, Ethiopia, were weak market links, poor infrastructure, underpayment of workers, and a growing sense of dependence. Therefore, measures should be taken to improve school infrastructure, establish strong relationships between producers and suppliers of SFP inputs and organizations, and have a budget that considers inflation and compensation for SFP workers during the summer into account. To decrease the perception of dependence on SFP among students and their families, efforts to raise awareness should also be implemented. According to the researchers, future studies should be needed, to quantify the perceived benefits and drawbacks of SFP to make it straightforward for policymakers and program designers to prioritize problems according to their magnitude.

## Supporting information

## Data Availability

The data used during the current study are available from the corresponding author upon request

## Acknowledgments

The manuscript is based on the doctoral research of Y.T.S. The Center for Food Science and Nutrition, Addis Ababa University, and Addis Ababa Education Bureau are acknowledged for facilitating the study. Last but not least, we are grateful to all the research participants for delivering the crucial information and thoughts necessary to accomplish this research.

## Author Contributions

***Conceptualization:*** Yihalem Tamiru

***Data curation:*** Yihalem Tamiru, Elyas Melaku

***Formal analysis:*** Yihalem Tamiru, Elyas Melaku

***Funding acquisition:*** Yihalem Tamiru

***Investigation:*** Yihalem Tamiru, Elyas Melaku

***Methodology:*** Yihalem Tamiru Semegn, Samson Gebremedhin Gebreselassie, Afework Mulugeta Bezabih, Abebe Ayelign Beyene, Elyas Melaku Mazengia.

***Project administration:*** Yihalem Tamiru Semegn, Samson Gebremedhin Gebreselassie, Afework Mulugeta Bezabih, Abebe Ayelign Beyene

***Resources:*** Yihalem Tamiru Semegn, Samson Gebremedhin Gebreselassie

***Supervision:*** Yihalem Tamiru Semegn, Samson Gebremedhin Gebreselassie, Afework Mulugeta Bezabih, Abebe Ayelign Beyene

***Visualization:*** Yihalem Tamiru Semegn, Samson Gebremedhin Gebreselassie, Afework Mulugeta Bezabih, Abebe Ayelign Beyene

***Writing original draft:*** Yihalem Tamiru Semegn

***Writing review & editing:*** Yihalem Tamiru Semegn, Samson Gebremedhin Gebreselassie, Afework Mulugeta Bezabih, Abebe Ayelign Beyene

***Ethical Standards Disclosure:*** The Institutional Review Board (IRB) of the College of Natural and Computational Sciences (CNS-IRB), Addis Ababa University, with Ref. No. CNCSDO/623/15/2023, approved all procedures involving research study participants by the Declaration of Helsinki. The Addis Ababa Education Bureau gave their permission. School directors and parents of participating students provided written informed consent before the study could start. Students took an active part in the study and provided their informed consent. All investigators, managers, and data gatherers adhered to strict confidentiality requirements.

## References

1. World Food Programme (WFP). State of School Feeding Worldwide 2022. Rome; 2022.

2. Kristjansson EA, Gelli A, Welch V, Greenhalgh T, Liberato S, Francis D, et al. Costs, and cost-outcome of school feeding programs and feeding programs for young children. Evidence and recommendations. Int J Educ Dev [Internet]. 2016;48:79–83. Available from: 10.1016/j.ijedudev.2015.11.011

3. Lesley Drake Alice Woolnough Imperial College London U, Burbano C, World Food Programme IDB, World Bank Group U, editors. Global School Feeding Sourcebook Lessons from 14 countries. 9HE, Imperial College Press 57 Shelton Street Covent Garden London WC2H; 2016.

4. Kretschmer A, Spinler S, Van Wassenhove LN. A school feeding supply chain framework: Critical factors for sustainable program design. Prod Oper Manag. 2014;23(6):990–1001.

5. Nkhoma OWW, Duffy ME, Cory-Slechta DA, Davidson PW, Mcsorley EM, Strain JJ, et al. Early-Stage Primary School Children Attending a School in the Malawian School Feeding Program Have Better Reversal Learning and Lean Muscle Mass Growth Than Those Attending a Non-SFP School. J Nutr. 2013;143:1324–30.

6. Wang D, Shinde S, Young T, Fawzi WW. Impacts of school feeding on educational and health outcomes of school-age children and adolescents in low-and middle-income countries: A systematic review and meta-analysis. J Glob Health. 2021;11:1–27.

7. Jomaa LH, Mcdonnell E, Probart C. School feeding programs in developing countries : impacts on children’s health and educational outcomes. Nutr Rev. 2011;69(2):83–98.

8. Donald Bundy, Carmen Burbano, Margaret Grosh, Aulo Gelli, Matthew Jukes, and LD. Rethinking School Feeding Social Safety Nets, Child Development, and the Education Sector. 2009.

9. Global FoodBanking Network GCNF. Developing a School Feeding Program. 2021.

10. Change MsTCGS of D and School Feeding: Interlocking of Planned Intervention and Lifeworlds of the Intervened. Wageningen University – Department of Social Sciences; 2016.

11. Destaw Z, Wencheko E, Zemenfeskidus S, Challa Y, Tiruneh M, Fite MT, et al. Use of modified composite index of anthropometric failure and MUAC-for-age to assess the prevalence of malnutrition among school-age children and adolescents involved in the school feeding program in Addis Ababa, Ethiopia. BMC Nutr. 2021;7(1):1–11.

12. Hussein K, Mekonnen TC, Hussien FM, Alene TD, Abebe MS. School Feeding and Nutritional Status of Students in Dubti District, Afar, Northeast Ethiopia: Comparative Cross-Sectional Study. Pediatr Heal Med Ther. 2023;Volume 14(June):217–30.

13. ICF CSA [Ethiopia]. Ethiopia Demographic and Health Survey. Addis Ababa, Ethiopia, and Rockville, Maryland, USA: CSA and ICF.; 2016.

14. Plaut D, Thomas M, Hill T, Worthington J, Fernandes M BNG to education outcomes: reviewing evidence from health and education interventions. Child and Adolescent Health and Development. 3rd Editio.

15. MoE. Education Statistics Annual Abstract: September 2019-March 2020 [Internet]. 2020. Available from: www.moe.gov.et

16. Mcewan PJ. The impact of Chile’s school feeding program on education outcomes. Econ Educ Rev [Internet]. 2013;32:122–39. Available from: 10.1016/j.econedurev.2012.08.006

17. Kristjansson B, Petticrew M, Macdonald B, Krasevec J, Janzen L, Greenhalgh T, et al. School feeding for improving the physical and psychosocial health of disadvantaged students (Review). Cochrane Database of Systematic Rev. 2009;(1).

18. Grantham-McGregor SM, Chang S, Walker SP, Al GET. Evaluation of school feeding programs: some Jamaican. Am J Clin Nutr. 1998;67.

19. Mary and Mbewe. An Exploration of the Views of Teachers, Pupils, and Parents Towards School Feeding Programme in Primary Schools in Ndola District : A Case for Kan’Gonga Area. Int J Multi-Disciplinary Res. 2018;1–81.

20. Weldeghebrael EH. Addis Ababa: City Scoping Study. 2021.

21. MoE. Federal Democratic Republic of Ethiopia, Ministry of Education Statistics Annual Abstract (ESAA) [Internet]. 2022. Available from: file:///C:/Users/HP/Downloads/ESAA 2014 EC (2021-22 G.C) Final.pdf%0Awebsite: www.moe.gov.et Email:

22. Baxter P, Jack S. Qualitative Case Study Methodology: Study Design and Implementation for Novice Researchers. Qual Rep. 2015;13(4):544–59.

23. Tong A, Sainsbury P, Craig J. Consolidated criteria for reporting qualitative research (COREQ): A 32-item checklist for interviews and focus groups. Int J Qual Heal Care. 2007;19(6):349–57.

24. Sim J, Saunders B, Waterfield J, Kingstone T. Can sample size in qualitative research be determined a priori? Int J Soc Res Methodol [Internet]. 2018;21(5):619–34. Available from: 10.1080/13645579.2018.1454643

25. Palinkas LA, Horwitz SM, Green CA, Wisdom JP, Duan N, Hoagwood K. Purposeful Sampling for Qualitative Data Collection and Analysis in Mixed Method Implementation Research. Adm Policy Ment Heal [Internet]. 2015;42(5):533–44. Available from: 10.1007/s10488-013-0528-y

26. Saunders B, Sim J, Kingstone T, Baker S, Waterfield J, Bartlam B, et al. Saturation in qualitative research: exploring its conceptualization and operationalization. Qual Quant. 2018;52(4):1893–907.

27. Akeju DO, Vidler M, Oladapo OT, Sawchuck D, Qureshi R, Von Dadelszen P, et al. Community perceptions of pre-eclampsia and eclampsia in Ogun State, Nigeria: A qualitative study. Reprod Health [Internet]. 2016;13(1). Available from: 10.1186/s12978-016-0134-z

28. Boene H, Vidler M, Sacoor C, Nhama A, Nhacolo A, Bique C, et al. Community perceptions of pre-eclampsia and eclampsia in southern Mozambique. Reprod Health [Internet]. 2016;13(1). Available from: 10.1186/s12978-016-0135-y

29. Braun V, Clarke V, Braun V, Clarke V. Applied Qualitative Research in Psychology. Appl Qual Res Psychol. 2017;0887(2006).

30. Kumpfer KL. The resilience framework. Resil Dev Posit Life Adapt. 1999;179–224.

31. Mohammed B, Belachew T, Kedir S, Abate KH. Effect of school feeding program on academic performance of primary school adolescents: A prospective cohort study. Clin Nutr ESPEN [Internet]. 2023;56:187–92. Available from: 10.1016/j.clnesp.2023.05.017

32. Meshkovska B, Gebremariam MK, Atukunda P, Iversen PO, Wandel M, Lien N. Barriers and facilitators to implementation of nutrition-related actions in school settings in low-and middle-income countries (LMICs): a qualitative systematic review using the Consolidated Framework for Implementation Research (CFIR). Implement Sci Commun. 2023;4(1):1–20.

33. Cammelbeeck S. Home Grown School Feeding Programme, Zambia. 2020;7(3):2017–22.

34. Agu CI, Ossai EN, Ogah OE, Agu IC, Akamike I, Ugwu GO, et al. An appraisal of the implementation of the national school feeding program and its effect on enrolment and attendance in public primary schools in Southeast, Nigeria: perception of heads of schools. BMC Nutr. 2023;9(1):1–10.

35. Desalegn TA, Gebremedhin S, Stoecker BJ. Successes and challenges of the Home-grown School Feeding Program in Sidama Region, Southern Ethiopia: a qualitative study. J Nutr Sci. 2022;11:1–7.

36. Philbrick SR. Families Experiences with Access to Universal Free School Meals During the COVID-19 Pandemic. Milligan University, Tennessee; 2023.

37. Jalal P, Sareen N. Perspectives of school children, teachers and parents towards school lunch program. J Res Humanit Soc Sci. 2022;10(12):400–6.

38. Education Ethiopia FM of. National School Feeding Policy Draft. 2019.

39. Fungo R. Implementation of the school feeding and nutrition programs in Uganda and the contribution of school meals to recommended dietary allowances (RDAs) of children : Challenges and opportunities. African J Food Sci. 2023;17(5):85–101.

40. Lemma M. The Practice and Challenges of School Feeding Program at Yenat Weg Charitable Society. St. Marys University; 2020.

